# Does Ethnicity Alter the Hazard of Stroke in Patients with Major Modifiable Cardiometabolic Risk Factors? A population-based Longitudinal Study using Electronic Health Records in South London

**DOI:** 10.1101/2024.04.08.24305017

**Authors:** Marc Delord, Abdel Douiri

## Abstract

**Background:** Ethnic inequality in stroke risk was partially explained by a history of hypertension, diabetes and socioeconomic status. We aimed to estimate the impact of ethnicity on the hazard of stroke in patients with hypertension, diabetes or hypercholesterolaemia in an urban, multiethnic, youthful and deprived population.

**Methods:** Multistate models were constructed using electronic health records, including the date of stroke onset, stroke risk factors, and the Index of Multiple Deprivation in Asian, Black or White adult patients registered at 41 general practices in South London between 2005 and 2021. Transitions to hypertension, diabetes or hypercholesterolaemia (transition 1), and from these cardiometabolic risk factors to stroke (transition 2) were considered.

**Results:** The study included 651,888 of the 849,968 registered patients (76.7%), of whom 8.6% were Asian, 18.2% Black, and 73.2% White, while 54.1% were female. Stroke incidence was 2.1% in Black patients, 1.2% in Asian patients, and 1.0% in White patients. Compared to White patients, Asian and Black patients had generally a higher risk of transitioning to hypertension, diabetes or hypercholesterolaemia (transition 1). Ethnicity however, did not affect transition to stroke from cardiometabolic risk factors (transition 2), except for hypercholesterolaemia, where Black patients had a higher risk of strokes.

**Conclusion:** Our results confirm that Black ethnicity does not confer excess hazard of stroke in patients with hypertension or diabetes. This observation was also made for Asian ethnicity. An excess hazard of stroke associated with Black ethnicity was observed only in patients with hypercholesterolaemia.

## Background

The excess stroke incidence in populations of Black compared to White ethnicity is well documented [1, 2, 3, 4, 5]. This excess has been associated with a higher prevalence of modifiable cardiometabolic stroke risk factors in Black patients, especially hypertension and diabetes [1]. However, the extent to which these risk factors account for this excess risk of stroke in Black patients remains controversial, even in studies reporting that the disparity in stroke incidence between Black and White populations could not be fully explained by hypertension and diabetes alone. Yet the addition of socioeconomic confounders such as income, education level or more general area-based measures like the Index of Multiple Deprivation (IMD), which captures broader contextual deprivation, generally provides further insight into observed inequalities in stroke risk and has been shown to be significantly associated with poorer outcomes [6, 3, 7]. Furthermore, discrepancies between studies suggest that alternative factors such as childhood socioeconomic status [3, 8], genetic or epigenetic factors [9, 10, 11], or even the contribution of traditional risk factors may still explain part of this difference.

Data on the incidence of stroke and the prevalence of associated risk factors among patients of Asian ethnicity are less abundant. Available studies report a higher incidence of stroke and risk factor profiles, similar to those observed in Black patients, characterised by a high prevalence of hypertension, cardiac diseases and notably diabetes. However, the sociodemographic profiles of these patients resemble those of White patients, with little social gradient reported between ethnic groups [12, 13].

The contribution of modifiable cardiometabolic risk factors such as hypertension, diabetes and hypercholesterolaemia to stroke risk across different ethnic backgrounds remains poorly understood. This question can be effectively addressed using population-based longitudinal electronic health records within the framework of multistate models [14]. Using this class of models, we aim to evaluate the impact of ethnicity on the hazard of three modifiable cardiometabolic risk factors (namely, hypertension, diabetes, and hypercholesterolaemia), and on the hazard of stroke mediated by these cardiometabolic risk factors, in a young, multiethnic, urban population.

## Methods

### Study participants

The study was conducted in the Lambeth borough of South London, a densely populated, multiethnic, and demographically youthful area. Approximately 40% of residents in Lambeth identify with ethnic groups other than White. The population aged 20 to 40 years is overrepresented in this Borough compared to both the London and UK averages. According to the 2019 Index of Multiple Deprivation (IMD), Lambeth ranks among the most socioeconomically deprived local authorities in England, with high levels of deprivation across multiple domains, including income, employment, education, housing, health, and the living environment [15, 16].

We utilised primary care electronic health records from adult patients of Asian, Black or White ethnicity, aged over 18, registered in any of the 41 Lambeth general practices between April 2005 and April 2021. A total of 3.2% of patients from the registry dropped out of the anonymised data-sharing scheme and were therefore excluded from the study. Restriction to patients of Asian, Black or White ethnicity was applied to ensure consistency with previous published results [1, 3, 6, 13, 17] and to ensure sufficiently large sample sizes within analysed subgroups.

### Primary care data

The data consisted of anonymised, patient-level information from ‘real-world’, routinely collected records, coded using Read codes and Systematized Nomenclature of Medicine – Clinical Terms (SNOMED – CT), and managed within the Egton Medical Information Systems (EMIS). Electronic health records included the date of diagnosis of 18 long-term conditions coded in accordance with the Quality and Outcomes Framework (QOF), based on QOF38 definitions. Anonymised preprocessed electronic health recods were provided by the Lambeth DataNet upon project approval by the Lambeth Clinical Commissioning Group. Long-term conditions included stroke, hypertension, diabetes, hypercholesterolaemia (total cholesterol > 5.0 mmol/L), atrial fibrillation, heart failure, and myocardial infarction. Other information included gender (self-ascribed), ethnicity (self-ascribed, including Asian, Black, mixed, other/unknown, and White ethnicities), ‘ever smoking’ status, and the Index of Multiple Deprivation (IMD) 2019 (from 1-most deprived to 5-least deprived). The IMD is developed by the Ministry of Housing, Communities & Local Government (MHCLG) in England. It integrates data across seven domains-income, employment, education, health, crime, housing, and the living environment-across small geographic areas to assess population needs and inform policy decisions aimed at addressing regional inequalities [18]. The study was restricted to Asian, Black, and White patients.

### Stroke risk factors

For consistency with previous studies [1, 3, 6] and subject on data availability, we have restricted our analysis to risk factors related to traditional risk factors, demographic variables, and a measure of socioeconomic status. These include age at mid follow-up, gender, hypertension, diabetes, hypercholesterolaemia, ‘ever smoking’ status, cardiovascular disease history (prior myocardial infarction, coronary heart disease, and heart failure), atrial fibrillation, and the IMD 2019.

### Multistate models and statistical analyses

The multistate models used in this study are extensions of survival models, allowing the analysis of multiple time-to-event variables in association with covariates. This provides a more focused insight into the biological and clinical processes leading to disease by evaluating the impact of prognostic factors at different phases of disease progression. Multistate models have been applied in secondary analyses of clinical trials to better understand complex disease processes [19] or life course transitions from a healthy state to chronic diseases and death [20]. Other examples include their use in epidemiological settings, such as assessing the impact of social inequalities on transitions from health to multimorbidity, frailty, or disability to mortality [21]. Unlike applications in clinical trials, which focus on treatment effects and disease progression in controlled environments, this study examines real-world population health and the influence of socioeconomic factors on health transitions. It uses a progressive illness/death model, where intermediate events, such as the occurrence of one or more long-term conditions, can influence the probability of subsequent events, leveraging electronic health records.

The progressive illness / death multistate model can be adapted to the setting of stroke, where a cardiometabolic risk factor represents an intermediate state that patients may enter before eventually experiencing stroke. This progressive risk factor / disease model can be enriched by adding death as a semi-competing event for stroke and other relevant transitions, as shown in Figure 1.

**Figure 1:**
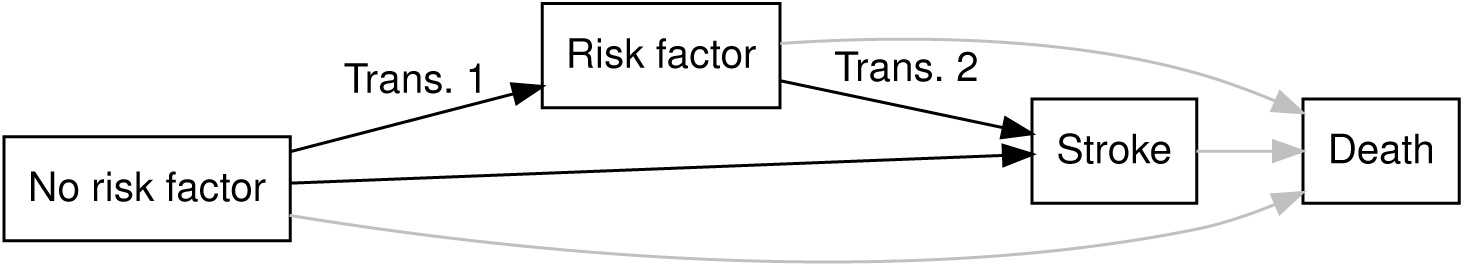
Progressive risk factor/disease model for stroke: The progressive risk factor / disease model is represented by black arrows. Grey arrows represent other estimable cause specific hazards

Three states are considered in this model. The first represents a state free from both the analysed risk factor and stroke. This state is represented by a rectangle labeled ‘No risk factor’. Patients diagnosed with the analysed risk factor transition to the state labeled ‘Risk factor’. Finally, upon experiencing stroke, patients enter the state labeled ‘Stroke’.

Arrows between the states represent possible transitions. In our study, transition 1 refers to the transition from the ‘No risk factor’ state to the ‘Risk factor’ state, while transition 2 refers to the transition from the ‘Risk factor’ state to the ‘Stroke’ state. The intensity of transitions between states can be estimated within the framework of a time-to-event regression model.

In this study, we analysed hypertension, diabetes, and hypercholesterolaemia sequentially as risk factors associated with stroke, as shown in Figure 1. Transition-specific hazard ratios (HR) were computed using a Cox proportional hazards model stratified by transitions and adjusted for all other risk factors as mentioned above. We restricted our analyses to the impact of ethnicity on transitions 1 and 2, i.e., the transition from the absence of the considered risk factor and the transition from the analysed risk factor to the record of stroke, respectively.

Multi-state models assume the Markovian assumption, which states that the future depends on the history only through the present state. However, for a more realistic representation of patients’ history, this assumption can be relaxed in several ways. The approach used in this study leads to state-arrival extended semi-Markov models in which the transition hazard may depend on the time of arrival in the present state. This allows for instance, to account for patients’ age at record of considered risk factors when estimating transition hazards from the risk factors states to stroke. State-arrival extended semi-Markov were constructed by adding patients’ age at entry of the risk factor state when estimating the hazard of transition to stroke from the risk factor state [22]. Of note, the proposed method focuses on transition hazards between possibly competing states (stroke or death). This aligns with our objective, which is to estimate the impact of covariates on transition rates, which is better addressed using cause-specific hazard models [23].

Long-term conditions, sociodemographic variables, and other variables were presented as frequencies or as median and interquartile range, as appropriate. The cumulative incidence of cardiometabolic stroke risk factors was estimated using the Kalbfleisch and Prentice method [24] versus death and stroke. All analyses were conducted on patients with complete covariates, as appropriate.

All computations were performed using the R language and environment for statistical computing (version 4.3.0 (2023-04-21)) [25]. More specifically, the following libraries were used: dplyr and ggplot2 (for data manipulation and visualisation, respectively) [26], mstate (for multistate model data preparation) [27], survival (for the Cox proportional hazards model) [28], and cmprsk (for the computation of cumulative incidence) [29].

### Ethics

#### Patient and Public Involvement

Since this research is based on secondary data analysis, there was no direct patient involvement in the design of the study, the development of research questions, or the selection of outcome measures. However, stakeholder consultations were conducted with Patient and Public Involvement (PPI) groups (Lambeth DataNet engagement groups) and general practitioner practice managers to develop materials informing patients about Lambeth DataNet’s purpose and data usage [30]. Accordingly, the results of this research will be disseminated widely. Efforts will include sharing findings with the public through accessible platforms, such as community websites and publications, as recommended by the Lambeth DataNet engagement groups. The results will also be communicated to local community groups, healthcare professionals, and PPI groups to facilitate further discussion and identify new areas of research.

## Results

### Patient characteristics

The study included 651,888 of the 849,968 registered patients (76.7%), of whom 8.6% were Asian, 18.2% Black, and 73.2% White, while 54.1% were female. Gender was not declared by 15 patients. The IMD was available for 643,317 (98.7%) patients. White and Asian patients had similar IMD profiles, whereas Black patients were overrepresented in IMD quintiles 1 and 2 (most deprived). Black patients had the highest rate of incident stroke (2.1%) as well as risk factors, including heart failure (1.3%), hypertension (19.7%), hypercholesterolaemia (30.6%), peripheral arterial/vascular disease (0.5%), and diabetes (10.4%). White patients had the lowest rate of incident stroke (1.0%) as well as coronary heart disease (1.2%), heart failure (0.7%), hypertension (6.6%), hypercholesterolaemia (19.0%), and diabetes (2.8%). Among Asian patients, rates were 1.2% for stroke, 10.0% for hypertension, 24.7% for hypercholesterolaemia, 8.5% for diabetes, 2.1% for coronary heart disease and 1% for myocardial infarction (table 1). The median follow up was 17.1 (Inter quartile range: from 11.2 to 29.1). Figure 2 summarises patients selection workflow, with comprehensive data available for 643,303 patients (98.7%).

**Figure 2:**
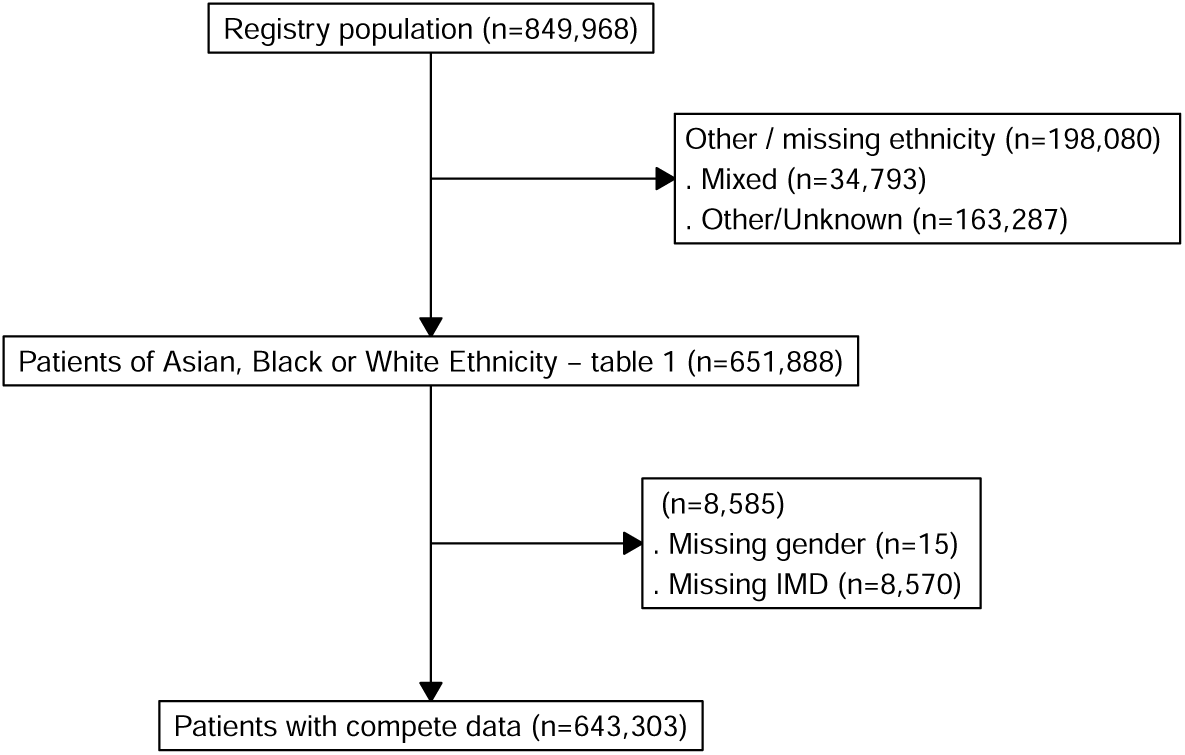
Diagram of patient selection

**Table 1:** Sociodemographics and stroke risk factors across ethnic groups.

| Ethnicity |  | Asian |  |  | Black |  |  | White |  |  | Total |
| --- | --- | --- | --- | --- | --- | --- | --- | --- | --- | --- | --- |
|  |  | No stroke | Stroke | Sub-total | No stroke | Stroke | Sub-total | No stroke | Stroke | Sub-total |  |
| Total N (%) |  | 55417 (98.8) | 645 (1.2) | 56062 (8.6) | 116439 (97.9) | 2495 (2.1) | 118934 (18.24) | 471893 (99.0) | 4999 (1.0) | 476892 (73.16) | 651888 |
| Age at end of follow-up | Median (IQR) | 35.2 (18.3) | 76.1 (20.1) | 35.5 (18.6) | 42.1 (24.0) | 72.1 (23.0) | 42.1 (24.8) | 34.1 (15.0) | 75.9 (24.0) | 34.1 (15.0) | 35.1 (17.9) |
| Sex | Male | 25459 (45.9) | 368 (57.1) | 25827 (46.1) | 54626 (46.9) | 1310 (52.5) | 55936 (47.0) | 214811 (45.5) | 2695 (53.9) | 217506 (45.6) | 299269 (45.9) |
|  | Female | 29958 (54.1) | 277 (42.9) | 30235 (53.9) | 61813 (53.1) | 1185 (47.5) | 62998 (53.0) | 257067 (54.5) | 2304 (46.1) | 259371 (54.4) | 352604 (54.1) |
| IMD<sup>a</sup> | 1 (most deprived) | 7295 (13.4) | 105 (16.4) | 7400 (13.5) | 33957 (29.5) | 758 (30.6) | 34715 (29.5) | 64970 (14.0) | 1117 (22.5) | 66087 (14.0) | 108202 (16.8) |
|  | 2 | 24319 (44.8) | 289 (45.2) | 24608 (44.8) | 56405 (48.9) | 1169 (47.1) | 57574 (48.9) | 217833 (46.8) | 2112 (42.6) | 219945 (46.7) | 302127 (47.0) |
|  | 3 | 15377 (28.3) | 178 (27.9) | 15555 (28.3) | 20833 (18.1) | 467 (18.8) | 21300 (18.1) | 136289 (29.3) | 1347 (27.2) | 137636 (29.2) | 174491 (27.1) |
|  | 4 | 6596 (12.1) | 60 (9.4) | 6656 (12.1) | 3636 (3.2) | 78 (3.1) | 3714 (3.2) | 36634 (7.9) | 327 (6.6) | 36961 (7.9) | 47331 (7.4) |
|  | 5 (least deprived) | 752 (1.4) | 7 (1.1) | 759 (1.4) | 407 (0.4) | 9 (0.4) | 416 (0.4) | 9936 (2.1) | 55 (1.1) | 9991 (2.1) | 11166 (1.7) |
| Hypertension | No | 50197 (90.6) | 279 (43.3) | 50476 (90.0) | 94449 (81.1) | 997 (40.0) | 95446 (80.3) | 442698 (93.8) | 2832 (56.7) | 445530 (93.4) | 591452 (90.7) |
|  | Yes | 5220 (9.4) | 366 (56.7) | 5586 (10.0) | 21990 (18.9) | 1498 (60.0) | 23488 (19.7) | 29195 (6.2) | 2167 (43.3) | 31362 (6.6) | 60436 (9.3) |
| Diabetes | No | 50898 (91.8) | 414 (64.2) | 51312 (91.5) | 104759 (90.0) | 1760 (70.5) | 106519 (89.6) | 459319 (97.3) | 4312 (86.3) | 463631 (97.2) | 621462 (95.3) |
|  | Yes | 4519 (8.2) | 231 (35.8) | 4750 (8.5) | 11680 (10.0) | 735 (29.5) | 12415 (10.4) | 12574 (2.7) | 687 (13.7) | 13261 (2.8) | 30426 (4.7) |
| Hypercholesterolaemia | No | 41872 (75.6) | 319 (49.5) | 42191 (75.3) | 81231 (69.8) | 1328 (53.2) | 82559 (69.4) | 383799 (81.3) | 2700 (54.0) | 386499 (81.0) | 511249 (78.4) |
|  | Yes | 13545 (24.4) | 326 (50.5) | 13871 (24.7) | 35208 (30.2) | 1167 (46.8) | 36375 (30.6) | 88094 (18.7) | 2299 (46.0) | 90393 (19.0) | 140639 (21.6) |
| Coronary Heart Disease | No | 54373 (98.1) | 518 (80.3) | 54891 (97.9) | 115064 (98.8) | 2303 (92.3) | 117367 (98.7) | 466566 (98.9) | 4387 (87.8) | 470953 (98.8) | 643211 (98.7) |
|  | Yes | 1044 (1.9) | 127 (19.7) | 1171 (2.1) | 1375 (1.2) | 192 (7.7) | 1567 (1.3) | 5327 (1.1) | 612 (12.2) | 5939 (1.2) | 8677 (1.3) |
| Heart Failure | No | 55011 (99.3) | 606 (94.0) | 55617 (99.2) | 115099 (98.8) | 2344 (93.9) | 117443 (98.7) | 468707 (99.3) | 4759 (95.2) | 473466 (99.3) | 646526 (99.2) |
|  | Yes | 406 (0.7) | 39 (6.0) | 445 (0.8) | 1340 (1.2) | 151 (6.1) | 1491 (1.3) | 3186 (0.7) | 240 (4.8) | 3426 (0.7) | 5362 (0.8) |
| PA/VD<sup>b</sup> | No | 55249 (99.7) | 619 (96.0) | 55868 (99.7) | 115934 (99.6) | 2429 (97.4) | 118363 (99.5) | 470014 (99.6) | 4799 (96.0) | 474813 (99.6) | 649044 (99.6) |
|  | Yes | 168 (0.3) | 26 (4.0) | 194 (0.3) | 505 (0.4) | 66 (2.6) | 571 (0.5) | 1879 (0.4) | 200 (4.0) | 2079 (0.4) | 2844 (0.4) |
| Myocardial infarction | No | 54924 (99.1) | 591 (91.6) | 55515 (99.0) | 115815 (99.5) | 2403 (96.3) | 118218 (99.4) | 469064 (99.4) | 4654 (93.1) | 473718 (99.3) | 647451 (99.3) |
|  | Yes | 493 (0.9) | 54 (8.4) | 547 (1.0) | 624 (0.5) | 92 (3.7) | 716 (0.6) | 2829 (0.6) | 345 (6.9) | 3174 (0.7) | 4437 (0.7) |
| Atrial Fibrillation | No | 55122 (99.5) | 590 (91.5) | 55712 (99.4) | 115543 (99.2) | 2373 (95.1) | 117916 (99.1) | 466852 (98.9) | 4429 (88.6) | 471281 (98.8) | 644909 (98.9) |
|  | Yes | 295 (0.5) | 55 (8.5) | 350 (0.6) | 896 (0.8) | 122 (4.9) | 1018 (0.9) | 5041 (1.1) | 570 (11.4) | 5611 (1.2) | 6979 (1.1) |
| Ever Smoked | No | 39004 (70.4) | 441 (68.4) | 39445 (70.4) | 76172 (65.4) | 1504 (60.3) | 77676 (65.3) | 244607 (51.8) | 2343 (46.9) | 246950 (51.8) | 364071 (55.8) |
|  | Yes | 16413 (29.6) | 204 (31.6) | 16617 (29.6) | 40267 (34.6) | 991 (39.7) | 41258 (34.7) | 227286 (48.2) | 2656 (53.1) | 229942 (48.2) | 287817 (44.2) |

<sup>a</sup> Index of multiple deprivation
<sup>b</sup> Peripheral Arterial/Vascular Disease

Figure 3 displays the cumulative incidence of hypertension, diabetes, and hypercholesterolaemia (transition 1) across ethnicity from 18 to 90 years. Black, Asian, and White patients had, respectively, higher, intermediate, and lower cumulative incidences of hypertension. Both Asian and Black patients were associated with a higher cumulative incidence of diabetes compared to White patients. Finally, while the cumulative incidence of hypercholesterolaemia showed less variation between ethnicities, it was notably higher in the Asian population, particularly between the ages of 30 and 50.

**Figure 3:**
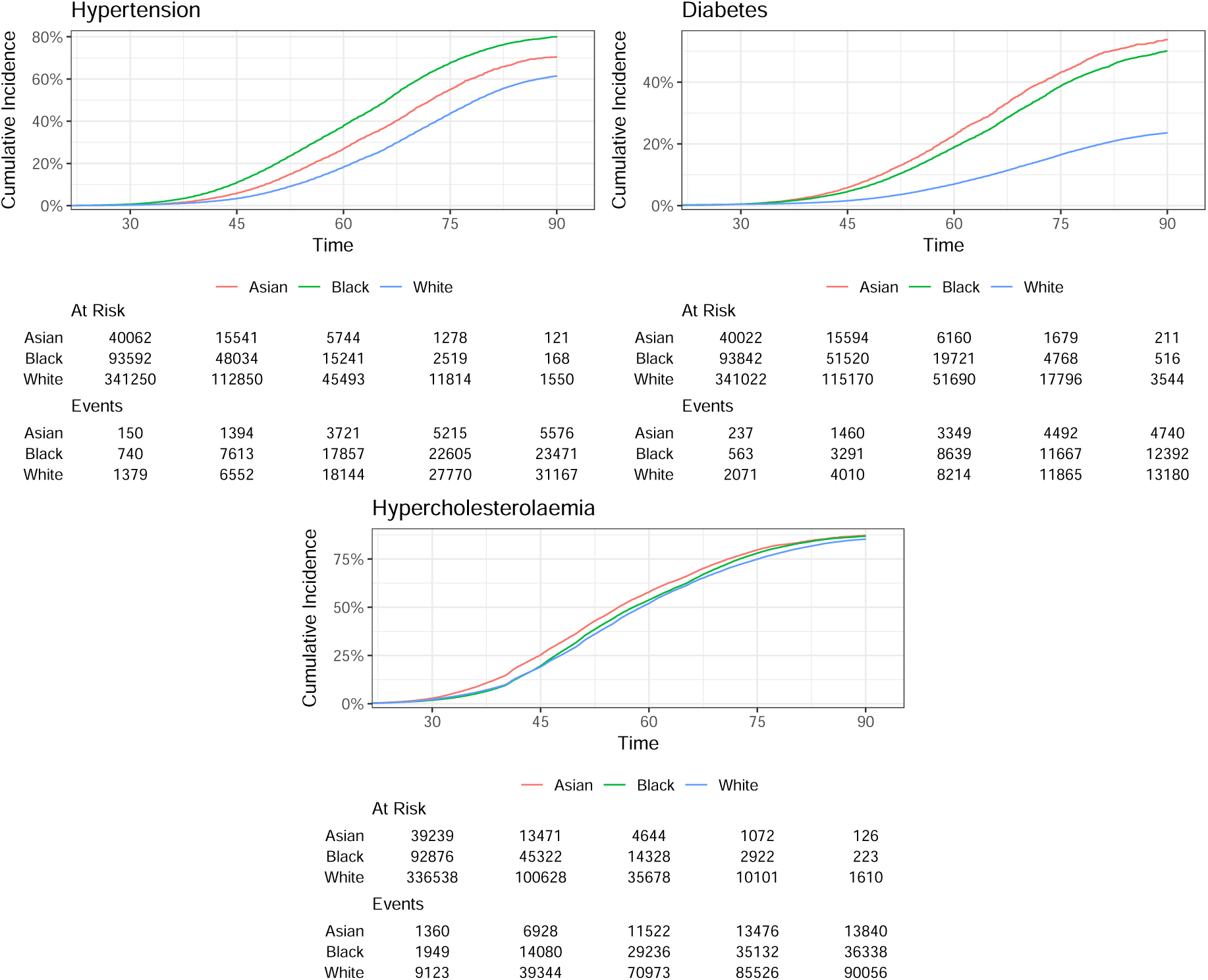
Cumulative incidence of hypertension, diabetes and hypercholesterolaemia across ethnicities between 18 and 90 years

### Multi-state model

Table 1 provides the number of patients transitioning from an absence of record of a given risk factor — including cardiometabolic risk factors — to the first recorded instance of that risk factor (transition 1, when transitioning to to hypertension, diabetes or hypercholesterolaemia), as well as the number of patients transitioning from a given risk factor to the first record of stroke (transition 2, when transitioning from hypertension, diabetes of hypercholesterolaemia). For example, 5,586 patients of Asian ethnicity transitioned from no record of hypertension to a first record of hypertension (10%). Among them, 366 (6.6%) subsequently had a record of stroke. Likewise, among Asian patients with no record of hypertension (n=50,476), 279 (0.6%) had a record of stroke. Supplementary figure S1 displays histograms of age at transition 1 and 2.

Table 2 displays unadjusted hazard ratios associated with Black and Asian patients compared to White patients for transitions to hypertension, diabetes, and hypercholesterolaemia (transition 1) and transitions from these cardiometabolic risk factors to stroke (transition 2). Likewise, Figure 4 illustrates adjusted hazard ratios for Black and Asian ethnicity compared to White ethnicity for transitions 1 and 2. Adjusted models showed that ethnicity was significantly associated with transitions to all three risk factors, with the highest values associated with diabetes for Asian and Black patients (HR: 3.05, 95% confidence interval (CI): 2.95–3.15, p<0.001, and HR: 2.35, CI: 2.29–2.41, p<0.001, respectively). Results were were less pronounced for hypertension, with high to moderate HRs associated with Black and Asian ethnicity, respectively (HR: 2.03, CI: 2.00–2.07, p<0.001, and HR: 1.26, CI: 1.23–1.30, p<0.001, respectively). Although highly significant, HRs associated with hypercholesterolaemia were relatively moderate (HR: 1.20, CI: 1.18–1.22, p<0.001, for Asian ethnicity, and HR: 0.98, CI: 0.96–0.99, p<0.001, for Black ethnicity). Except for Black ethnicity associated with hypercholesterolaemia (HR: 1.15, CI: 1.08–1.24, p<0.001), ethnicity did not significantly affect the hazard of transitions from tested cardiometabolic risk factors to stroke. Corresponding unadjusted hazard ratios showed an overall similar pattern, except for transition 2 associated with hypertension, where both Black (HR: 1.13, CI: 1.06–1.21, p<0.001) and Asian ethnicity (HR: 1.16, CI: 1.04–1.30, p=0.007) had higher risk compared to White ethnicity (Table 2).

**Figure 4:**
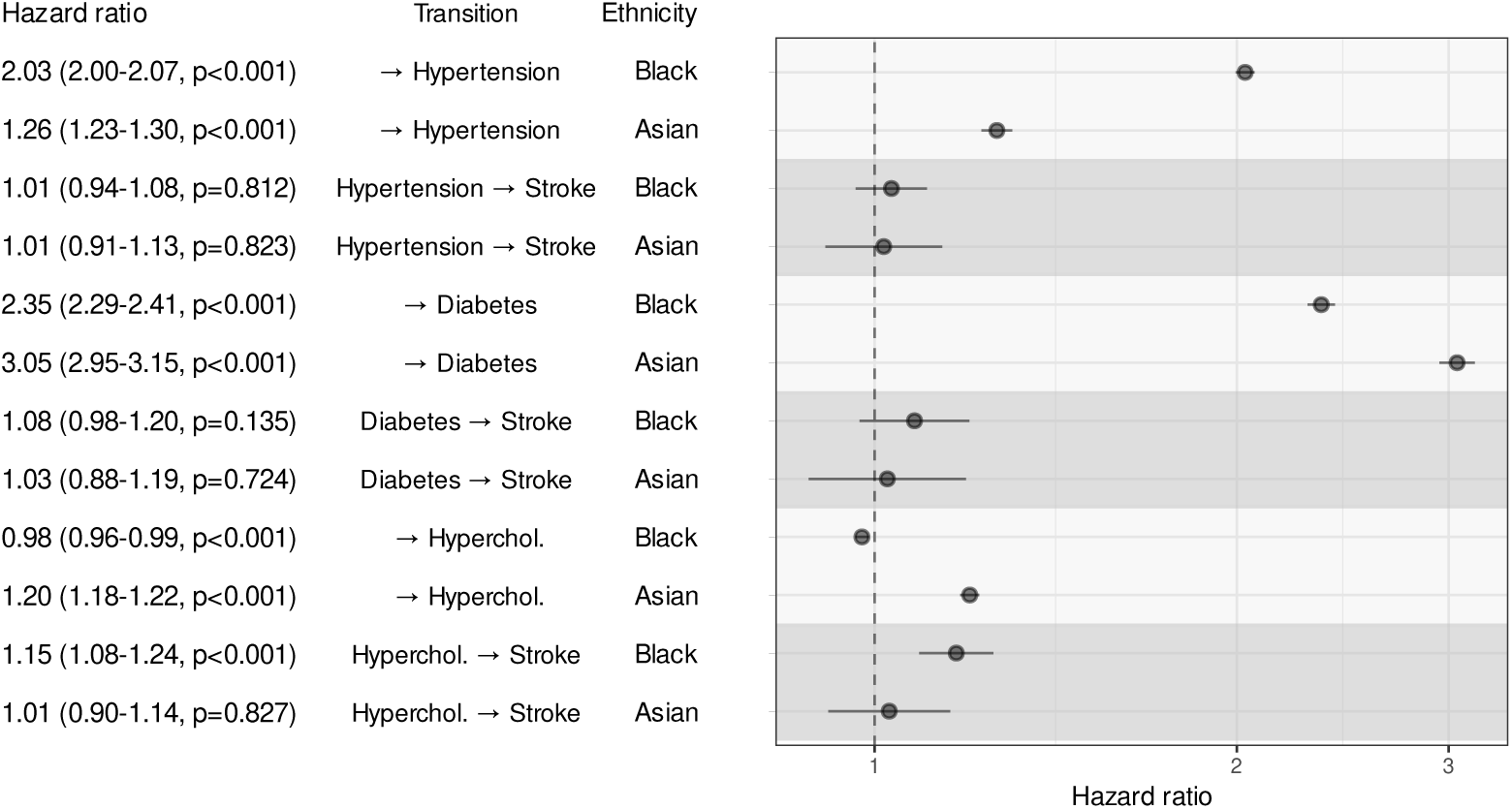
Adjusted transition-specific hazard ratios of the risk factor/disease models in which hypertension, diabetes and hypercholesterolaemia were sequentially analysed as risk factor associated with stroke. Hazard ratios are adjusted as appropriate for gender, hypertension, diabetes, hypercholesterolaemia, ‘ever smoking’ status, cardiovascular disease history (prior myocardial infarction, coronary heart disease, and heart failure), atrial fibrillation, and the IMD 2019. “→ Risk factor” indicates a transition from a state free from both the analysed risk factor and stroke, to the analysed risk factor (transition 1).

**Table 2:** Unadjusted transition-specific hazard ratios of the risk factor/disease models in which hypertension, diabetes and hypercholesterolaemia were sequentially analysed as risk factor associated with stroke. “→ Risk factor” indicates a transition from a state free from both the analysed risk factor and stroke, to the analysed risk factor (transition 1)

| Transition | Hazard Ratio | Ethnicity |
| --- | --- | --- |
| → Hypertension | 2.26 (2.22-2.30, $p<0.001$) | Black |
| → Hypertension | 1.43 (1.39-1.47, $p<0.001$) | Asian |
| Hypertension → Stroke | 1.13 (1.06-1.21, $p<0.001$) | Black |
| Hypertension → Stroke | 1.16 (1.04-1.30, $p=0.007$) | Asian |
| → Diabetes | 2.63 (2.56-2.69, $p<0.001$) | Black |
| → Diabetes | 3.04 (2.94-3.14, $p<0.001$) | Asian |
| Diabetes → Stroke | 1.08 (0.98-1.20, $p=0.128$) | Black |
| Diabetes → Stroke | 0.96 (0.83-1.12, $p=0.630$) | Asian |
| → Hyperchol. | 1.05 (1.04-1.07, $p<0.001$) | Black |
| → Hyperchol. | 1.21 (1.19-1.24, $p<0.001$) | Asian |
| Hyperchol. → Stroke | 1.25 (1.17-1.35, $p<0.001$) | Black |
| Hyperchol. → Stroke | 1.07 (0.95-1.20, $p=0.256$) | Asian |

## Discussion

This study set out to better understand the impact of ethnicity on the hazard of major modifiable cardiometabolic risk factors as well as on the hazard of stroke mediated by these risk factors in a multiethnic and urban population. Our findings do not support the hypothesis of excess hazard of stroke in Black patients with hypertension or diabetes. Similar observations were made in Asian patients, not only for hypertension and diabetes but also for hypercholesterolaemia. Our results also suggest that Black patients with hypercholesterolaemia have a higher hazard of stroke compared to their Asian or White counterparts.

Although previous studies have reported no excess risk of stroke in Black patients compared to White patients with hypertension or diabetes [1, 6], no clear consensus has emerged in the literature to date [3]. In line with previous findings, our results support the hypothesis of equipotency of hypertension and diabetes regarding stroke hazard in Black and White patients and bring new evidence that this hypothesis also holds for Asian patients. Our study confirms the role of hypertension and diabetes as major mediators of the excess incidence of stroke in Asian and Black patients [1, 3].

This excess incidence of stroke is attributable to the higher incidence and prevalence of these cardiometabolic risk factors in Asian and Black patients (figure 3 and table 1). Conversely, hypercholesterolaemia, which is more evenly distributed across ethnicities compared to hypertension and diabetes (table 1, figure 3 and 4), exhibits overall higher incidence rates (table 1 and figure 3). This establishes hypercholesterolaemia as a major mediator of stroke incidence, irrespective of ethnicity [31, 32].

Our findings also highlight the importance of hypercholesterolaemia in mediating the excess incidence of stroke in Asian and Black patients in two distinct ways: (1) in Asian patients, the excess incidence of stroke is directly mediated via the excess incidence of hypercholesterolaemia, mainly at younger ages (30–50 years) (Figure 3); and (2) in Black patients, the excess incidence of stroke is mainly due to a higher hazard of stroke in patients with hypercholesterolaemia (Figure 4).

Of note, the excess incidence of stroke mediated by modifiable cardiometabolic risk factors depends not only on the hazard of stroke associated with these risk factors, but also on their prevalence. Therefore, further investigation – beyond the scope of this study – is needed to better understand ethnic inequality in the incidence of stroke through: (i) evaluating the impact of observed ethnic disparity on the incidence of cardiometabolic risk factors (direct mediation); and (ii) evaluating the impact of excess hazard of stroke associated with ethnicity given cardiometabolic risk factors, such as with hypercholesterolaemia in our study (indirect mediation).

The absence of a significant ethnic-related excess of stroke hazard associated with hypertension and diabetes in our study, suggest that no other factor explains the excess in stroke incidence observed between Asian, Black and White patients. This further suggest the hypothesis of equal adherence and medical care in the observed population [1]. However, the excess hazard of stroke observed in Black patients with hypercholesterolaemia, may invalidate this hypothesis in this setting. Therefore, further research is needed to better evaluate the hypothesis of an excess hazard of stroke resulting from prescription disparities among Black patients with hypercholesterolemia compared to Asian and White patients [33].

Strengths and limitations of our study relate to the use of routinely collected primary care electronic health records, the population under study, or its limited geographical location. Though primary care electronic health records are considered a reliable source of data and have been increasingly used in research [34, 35], surveillance and public health planning [36], concerns may arise regarding accuracy or completeness of collected electronic health records [37]. For instance, information on current smoking status, the level of HDL and LDL or practice identitifyer was not available in electronic health records analysed in this study. Other potential explanatory factors not explored in our study include, for example, the levels and ratio of high- and low-density lipoprotein (HDL and LDL respectively) [38], body mass index [39], history of infectious diseases [40], the impact of mental health conditions [41], or more generally, the patterns of multimorbidity across different ethnic background [42, 43].

Some residual confounding may also persist, for instance, due to patterns in socioeconomic status or practice-level effects not fully captured by the IMD, or due to potential misclassification related to stroke ascertainment.

Stroke records in this study correspond to diagnoses as recorded by general practitioners according to standard clinical practice in the UK, which typically involves confirmation in secondary care through specialist assessment and brain imaging before the information is communicated back to primary care [44, 45], and coded using Read and SNOMED-CT terms in accordance with Quality and Outcomes Framework (QOF) definitions. However, as analysed records were anonymised, additional individual verification – including confirmation of stroke-free status at baseline – was not feasible. Consequently, some residual diagnostic misclassification can-not be excluded, even though stroke diagnoses in UK primary-care electronic health records largely reflect hospital-confirmed events and are considered reliable for epidemiological studies. [34, 35, 36].

Of note, both ethnicity and gender were self-ascribed variables, referring to individuals’ social and cultural identities, reflecting individuals’ sense of belonging, shaped by geographic, cultural, and sociopolitical factors rather than biological characteristics [46, 47]. If biological sex is relevant to health outcomes, gender is equally relevant, as it captures social and behavioural dimensions influencing health and care. In routine health records, these constructs often overlap substantially – over 99.95% in the UK Biobank, for instance [47] – and using self-ascribed gender represents a practical analytic choice reflecting this construct without presuming equivalence with biological sex.

On the other hand, analysis of longitudinal primary care electronic health records at the population scale enables the conduct of exploratory analysis allowing to characterise moderate differential trends such as the subtle ethnic inequality in the incidence of hypercholesterolaemia reported in our study. This approach offers insights into public health and epidemiology that may be more challenging to obtain through classical approaches, such as case-control studies or cohort studies [48, 49].

The characteristics of the analysed population are also important aspects to consider here, as they may hinder or favour particular study objectives. For instance, the relative socio-demographic uniformity within the Lambeth population tends to hinder the evaluation of the impact of the socio-economic status on cardiometabolic risk factors and stroke. However, this characteristic is not of prime importance in our study, as the socio-economic status is regarded as a confounder.

Likewise, the geographical unity of the study population is likely to mitigate the impact of differential environmental exposures, reducing the risk of unidentifiable confounding. In this context, the geographical and environmental homogeneity characterizing the study population can be regarded as a favorable aspect with regard to the addressed research question.

Finally, also in line with our study objectives, the multiethnic composition of the Lambeth population, particularly the substantial representation of Asian and Black patients, enhances its potential to serve as a valuable cohort for investigating ethnicity-related variations in health outcomes.

## Conclusion

Previous recommendations aiming to reduce inequality in stroke incidence between Black and White patients have focused on reducing ethnic disparities in the incidence of hypertension and diabetes [1, 3], which are major mediators of excess hazard of stroke in patients of Black ethnicity. Our findings align with these recommendations and provide new evidence to extend them to Asian patients, as well as to hypercholesterolaemia in these populations. Further research and alternative strategies will be needed to elucidate and address the excess hazard of stroke observed in Black patients with hypercholesterolaemia.

## Data Availability

Restrictions apply to the availability of the data that support the findings of this study, since they were used under an IRB approval, and hence not publicly available.

## List of abbreviations

SNOMED – CT: Systematized Nomenclature of Medicine – Clinical Terms
EMIS: Egton Medical Information Systems
QOF38: Quality and Outcomes Framework version 38 definitions
IMD: Index of Multiple Deprivation
MHCLG: Ministry of Housing, Communities & Local Government
PPI: Patient and Public Involvement
HR: Hazard Ratios
CI: Confidence Interval
HDL: High density Lipoprotein
LDL: Low Density Lipoprotein
NIHR: National Institute for Health and Care Research
HRA: Health Research Authority

## Declarations

### Ethics approval and consent to participate

This study was conducted in accordance with the Declaration of Helsinki. Data were provided by the Lambeth DataNet following approval for the analysis of fully anonymised data by the Lambeth Clinical Commissioning Group. All patients had been informed of potential secondary uses of their data through Fair Processing Notices and were given the opportunity to opt out [50]. Accordingly it was exempt from Research Ethics Committee review and the requirement for individual consent, in accordance with NHS Health Research Authority (HRA) guidance and relevant UK legislation, including the Data Protection Act 2018 (Chapter 2, Sections 8 and 10) and the Health and Social Care Act 2012 (Section 261).

### Consent for publication

Not applicable

### Availability of data and materials

Data supporting this study can be obtained from the Lambeth Datanet subject to IRB approval, and are not publicly available.

### Authors’ contributions

MD designed the study (conceptualisation and methodology); AD acquired the data; MD carried out data curation; MD and AD conducted investigations; MD developed computer code and performed formal analysis, including data visualisation and validation; MD and AD interpreted the data. MD drafted the manuscript, and AD supervised the research. MD and AD reviewed, edited, and approved the final version of the manuscript.

## Acknowledgments

We would like to express our deepest gratitude to Pr Mark Ashworth, who sadly passed away during the preparation of this manuscript. His contributions to this research were invaluable, and his dedication to the field continues to inspire us. This work is dedicated to his memory.

