## Supplementary materials for "Does Ethnicity Alter the Hazard of Stroke in Patients with Major Modifiable Cardiometabolic Risk Factors? A population-based Longitudinal Study using Electronic Health Records in South London"

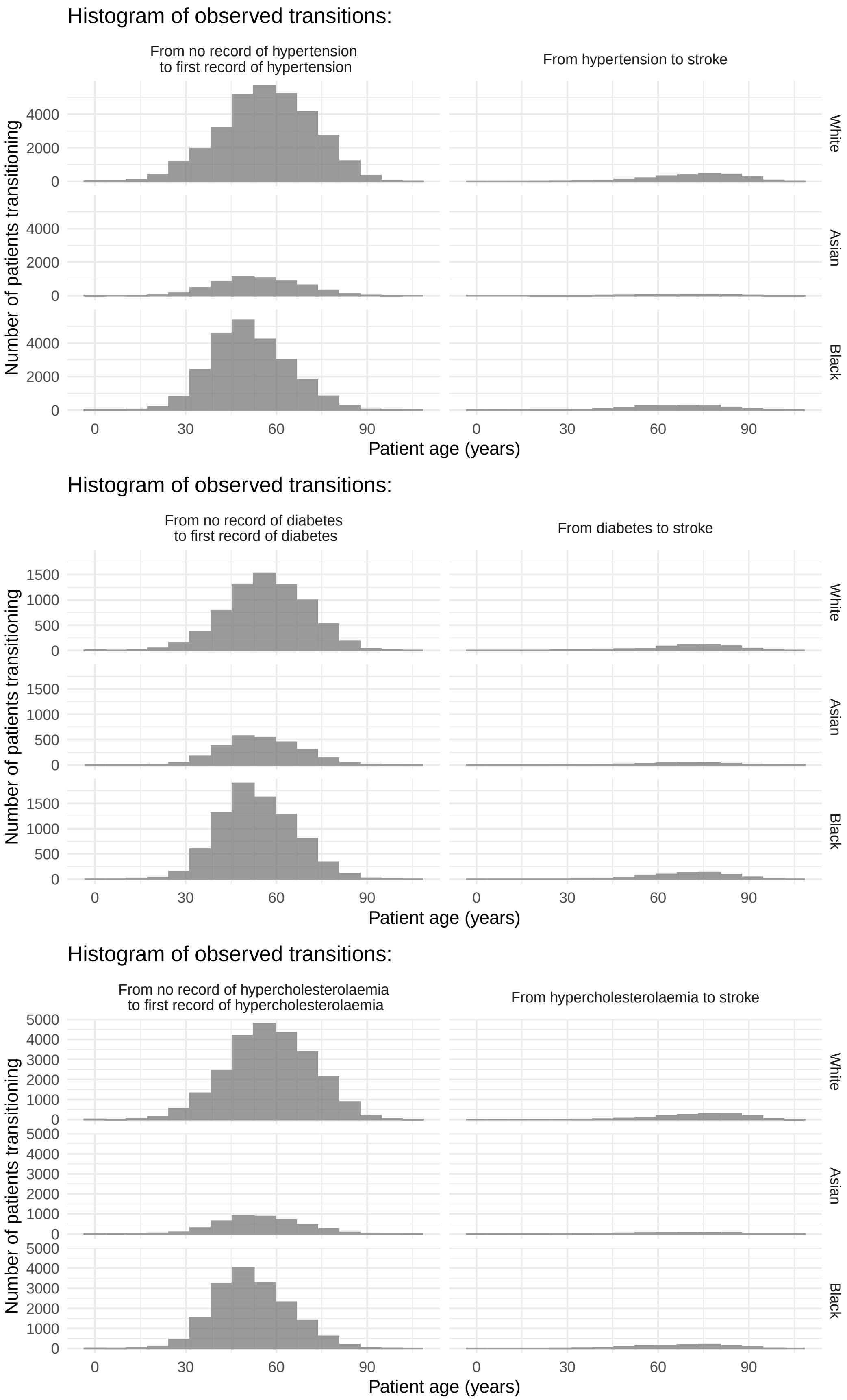


***Supplementary Figure S1:*** *Histograms of ages at transition 1 and 2: These graphs represent the number of patients and age at transition to the first record of the considered cardiometabolic in patient without a record of stroke and transition from the considered cardiometabolic risk factors to first record of stroke.*
